# Network-based biomarkers in background electroencephalography in childhood epilepsies – A scoping review and narrative synthesis

**DOI:** 10.1101/2024.05.17.24307531

**Authors:** Kay Meiklejohn, Leandro Junges, John R. Terry, Alison Whight, Rohit Shankar, Wessel Woldman

## Abstract

**Background:** Brain network analysis is an emerging field of research that could lead to the development, testing and validation of novel biomarkers for epilepsy. This could shorten the diagnostic uncertainty period, improve treatment, decrease seizure risk and lead to better management. This scoping review summarises the current state of electroencephalogram (EEG)-based network abnormalities for childhood epilepsies. The review assesses the overall robustness, potential generalizability, strengths, and limitations of the methodological frameworks of the identified research studies.

**Methods:** PRISMA guidelines for Scoping Reviews and the PICO framework was used to guide this review. Studies that evaluated candidate network-based features from EEG in children were retrieved from four international indexing databases (Cochrane Central / Embase / Medline / PsycINFO). Each selected study design, intervention characteristics, methodological design, potential limitations, and key findings were analysed.

**Results:** Of 2,959 studies retrieved nine were included. Studies used a group-level based comparison (e.g. based on a statistical test) or a classification-based method (e.g. based on a statistical model, such as a decision tree). A common limitation was the small sample-sizes (limiting further subgroup or confounder analysis) and the overall heterogeneity in epilepsy syndromes and age groups.

**Conclusion:** The heterogeneity of included studies (e.g. study design, statistical framework, outcome metrics) highlights the need for future studies to adhere to standardized frameworks (e.g. STARD) in order to develop standardized and robust methodologies. This would enable rigorous comparisons between studies, which is critical in assessing the potential of network-based approaches in developing novel biomarkers for childhood epilepsies.

## 1. Introduction

### 1.1 Background

Epilepsy is the most common chronic neurological condition of childhood, affecting over 60,000 children in the UK. Early intervention can improve seizure control and prevent deterioration of cognitive function [1]. However, diagnosis of epilepsy can be challenging and time intensive [2]. The primary diagnostic tool used in cases of suspected epilepsy is the electroencephalogram (EEG) [3]. In the absence of seizures, or other visible EEG abnormalities (such as interictal epileptiform discharges), confirming epilepsy is difficult [3]. Since these are unpredictable events for the majority of people with suspected seizures, time to diagnosis can be long (median: 1 year) [4], with high rates of misdiagnosis (>20%) [5]. Once diagnosed, people typically commence treatment with anti-seizure medications (ASMs). However, measuring response to ASMs is currently “watchful waiting”: periodically asking parents and carers whether treatment is effective from day-to-day observation [6]. Approximately a third (35%) never respond effectively, with timescales of over a year common for those who do [7]. Furthermore, a wide range of non-epileptic paroxysms can affect children [8], meaning paediatric misdiagnosis rates are likely greater [9]. Given the complexity and heterogeneity in epilepsy and seizures presentation and management, there is a clear need for novel epilepsy biomarkers that can more ably support the diagnosis, prognosis and management of epilepsy [10].

### 1.2 Rationale

During recent years, brain network disruptions or abnormalities modelling is being used to increasing understand epilepsy and seizures [11]. Functional network structures derived from modalities such as EEG have been studied in detail, with systematic reviews on generalised and focal epilepsies in [12,13] respectively. Although these reviews reported several studies with nonadult data only, there was limited focus on the impact of age on network abnormalities or dynamics.

No reviews were identified from publications or the PROSPERO database that provided an overview of network-based abnormalities from EEG specifically in the context of childhood epilepsies. A summary of the current state of research studies on network-based differences in the childhood epilepsies including assessments of the benefits and limitations (e.g. modality or methodological) and strengths and weaknesses could inform future directions for research studies.

### 1.3 Aim and research question

The aim of this scoping review was to identify studies which explored for candidate network-based biomarkers from waking, background clinical EEG in the childhood epilepsies. In particular, to identify specific network concepts that apply to 3 distinct childhood age groups (2-5 years, 5-10 years and 10-18 years). This review also looks for potential differences in the biomarkers between different epilepsy classifications and controls (e.g. healthy or a differential diagnosis).

## 2. Methods

### 2.1 Scope

The Preferred Reporting Items for Systematic Reviews and Meta-Analysis Extension for Scoping Reviews (PRISMA-ScR) and the Population, Intervention, Comparator, Outcome, and studies (PICO) frameworks were used to develop the search strategy structure the review (Table 1). Supplemental Material A provides the PRISMA-ScR framework [14].

**Table 1:**
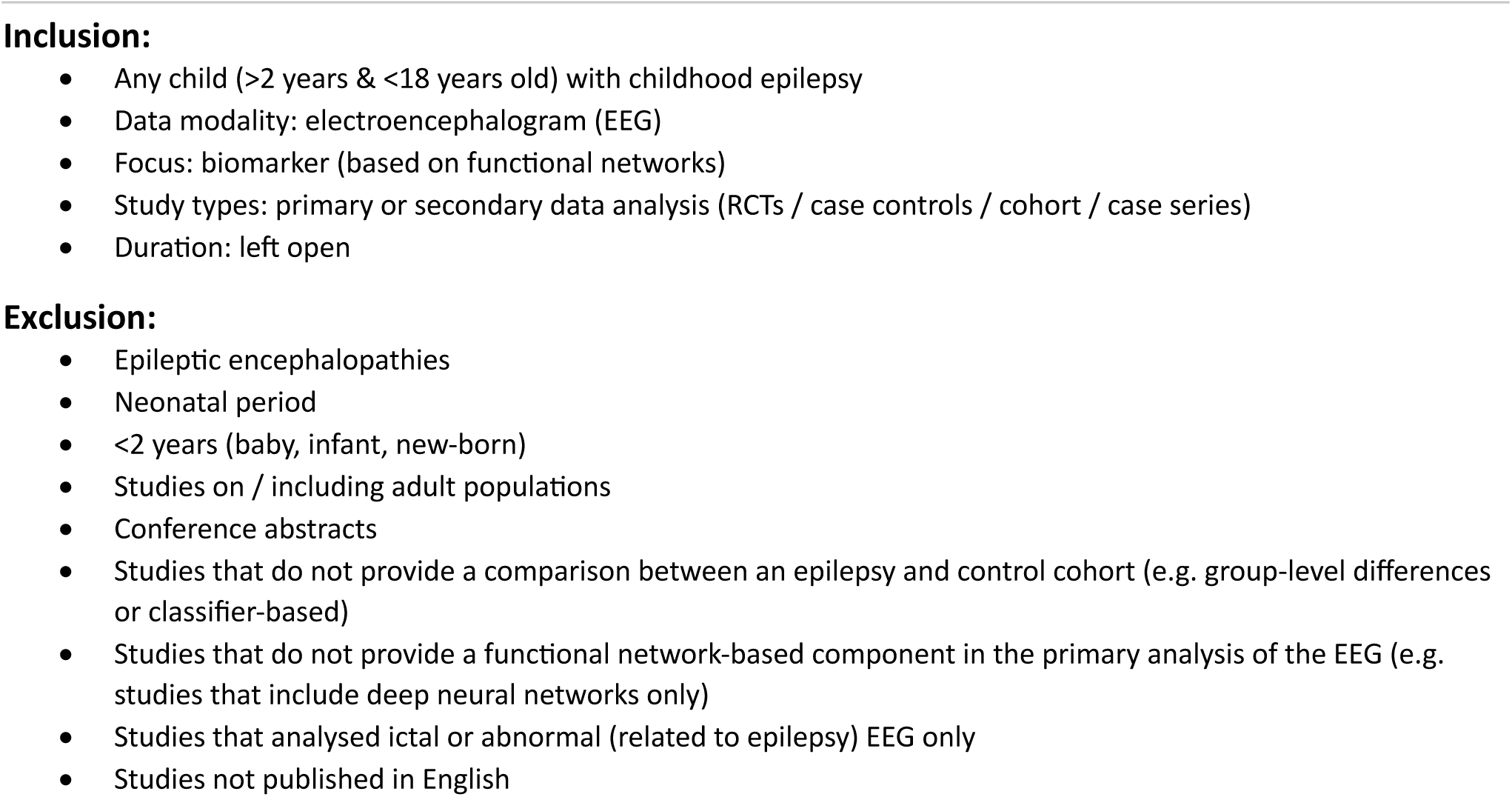
Inclusion and exclusion criteria Inclusion.

### 2.2 Eligibility criteria

The inclusion and exclusion criteria are summarised in Table 1. To identify candidate biomarkers relevant to paediatric populations, only studies that contained specific analysis on nonadult EEG data were eligible i.e. studies that conducted analysis on populations with ages ranging 14-30 years were ineligible if there was no secondary analysis made on children aged 14-18 years. However, the scope of the review was not limited to network-based markers only. If studies considered additional candidate biomarkers (e.g. spectral) these were also assessed and summarised. To provide a comprehensive summary of candidate network-based biomarkers, we did not restrict the type of EEG monitoring (e.g. high-density or intracranial are all eligible) but the chosen methodology in the paper would need to be appropriate for clinical EEG (as the most commonly used modality). Furthermore, since the scope of the review was to identify how these candidate features differ between epilepsy and control cohorts the majority of studies were expected to be based on scalp EEG. Although the focus of the review was on childhood epilepsies, the scope of the review was limited to that of subjects in the age range 2-18 years and those without epileptic encephalopathies. This conservative approach was chosen to reduce the inherent heterogeneity within the childhood epilepsies.

### 2.3 Search strategy

Four databases (Embase, Medline, PsycINFO and Cochrane Central) were searched on 19^th^ March 2024 by an information specialist (AW) with guidance from the authors to identify relevant candidate studies. Four key papers were hand selected as a validation test-set by two of the authors (LJ & WW) with extensive experience in the field of network-based biomarkers derived from EEG. The search string was structured based on 3 main themes Childhood Epilepsy (MeSH OR keywords) AND Network (MeSH OR keywords) AND Electroencephalogram (MeSH OR keywords). An example of the search string (with the corresponding number of results returned) is provided in the Supplemental Materials (B).

### 2.4 Screening and article selection

Identification of studies via 4 databases retrieved 2,959 records and these were uploaded to Rayyan via RIS files. This was done by an information specialist. Of this sample, 316 duplicates were identified and removed by one reviewer (KM). A sample (n=533) of the retrieved record were visually inspected by the same reviewer, via abstract only (focus was placed on keywords noted in section 2.3). Of these n=533, 80 were sought for retrieval and 453 were excluded. The 80 records sought for retrieval were identified as a “testing sample” to derive the best combination of keywords that would be used as Boolean framework to identify the remainder of the articles. Several combinations of keywords were assessed in combination (using Boolean search terms via the PICO framework) until a high retrieval rate of relevant papers (and key papers) were identified from the validation set (n=80).

The selected combination of Boolean keywords was then applied to the remaining articles (n=2110), retrieving n=55 papers that would then be combined with the previous n=80, totalling n=135 that would make full text review. Two reviewers (KM,WW) then independently reviewed n=135 full text articles. One reviewer (KM) selected nine articles for full review and the second reviewer (WW) selected the same nine articles plus two additional articles. A third reviewer (LJ) then reviewed the selected 11 articles, two were discarded at this point as they contained either source-based analysis on MEG or deep neural networks which were out of the scope of the study. Nine articles were left for full review, with all three reviewers in agreement for the final list.

### 2.5 Data extraction

Three reviewers then further appraised and extracted data from the final selection of studies (n=9). The PICO framework [15] was used to identify the key concepts of the topic and guide the authors on the information to be extracted, see Table 2.

**Figure 1:**
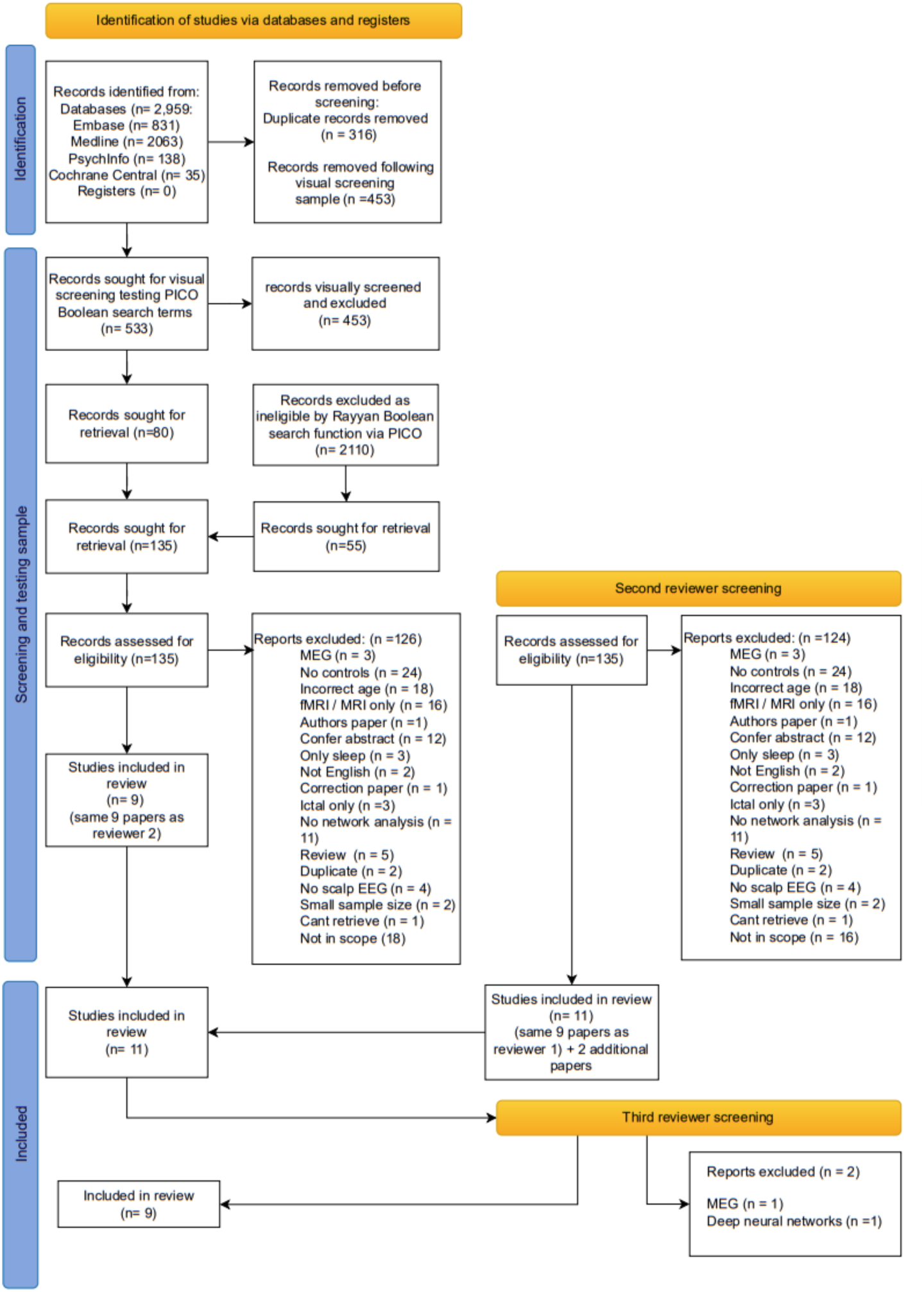
Article selection and reasons for exclusion (following the PRISMA guidelines)

**Table 2:** Data extraction.

| Article information | Data to be extracted |
| --- | --- |
| Study information | Year of publication<br>Epilepsy classification<br>Sample size (patients and seizures) |
| Intervention | Type of intervention<br>Description of intervention features / components<br>Recording details (e.g. equipment, technologist, montage)<br>Electrode details (e.g. placement, number, reference)<br>Sampling rate<br>Impedences (e.g. <5kΩ)<br>Patient state (e.g. maintenance of wakefulness, activations)<br>Pre-processing (e.g. filters, artefact removal, epoch selection)<br>Network analysis<br>Network derivation method (e.g. cross-correlation, PLI)<br>Network features (e.g. mean degree, clustering coefficient)<br>Statistical framework (e.g. cross-validation, multiple comparisons) |
| Evaluation | Main findings regarding network abnormalities (e.g. significant group-level differences, sensitivity, specificity, accuracy)<br>Strengths and weaknesses of the study |

### 2.6 Data analysis and synthesis

A scoping overview of the current state of the literature along with a narrative analysis of the data extracted from the research studies was conducted by three authors. This includes an assessment of the strengths and weaknesses (e.g. potential bias) of the studies identified.

## 3. Results

### 3.1 Search results

**Table 3.**
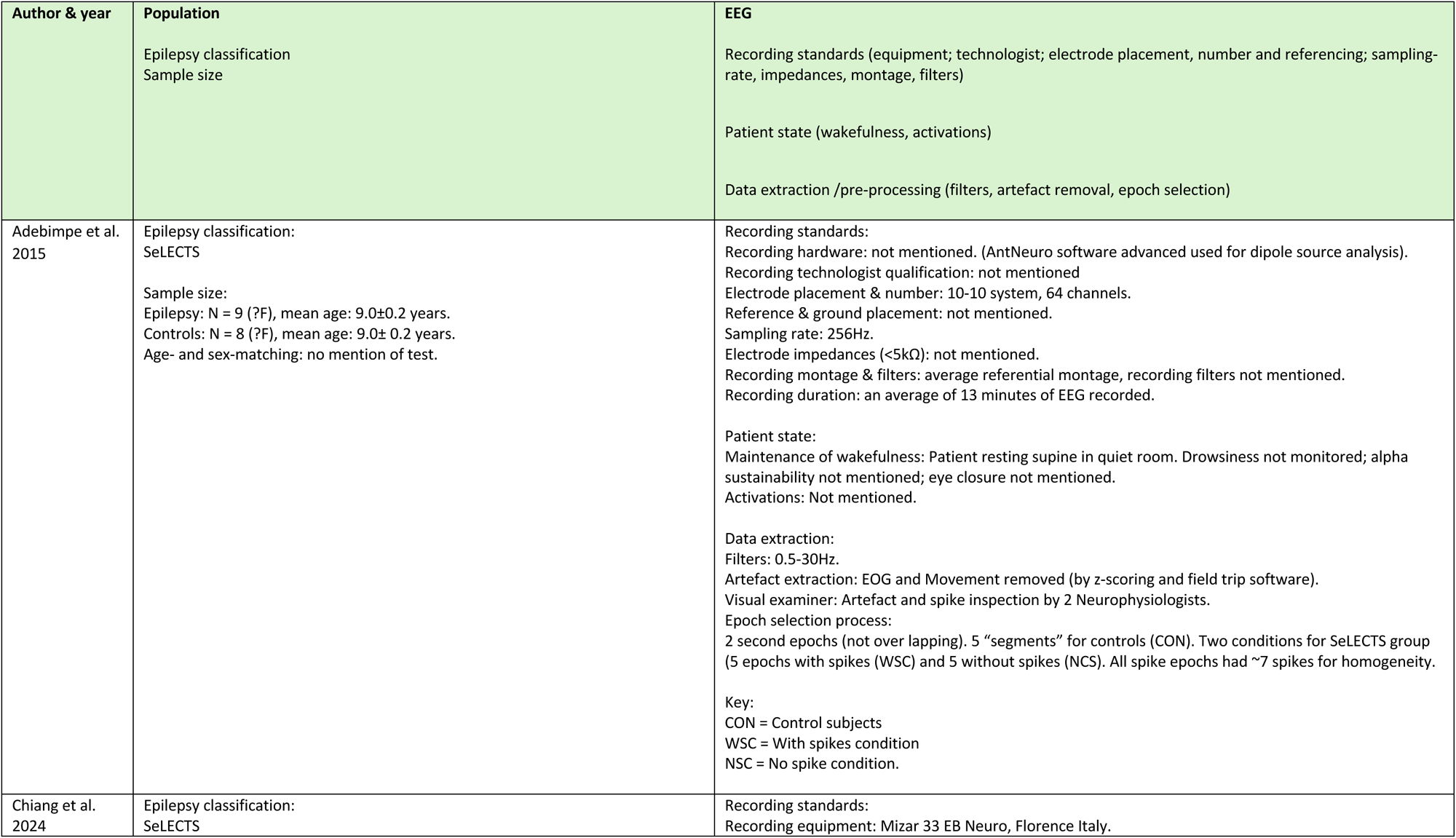
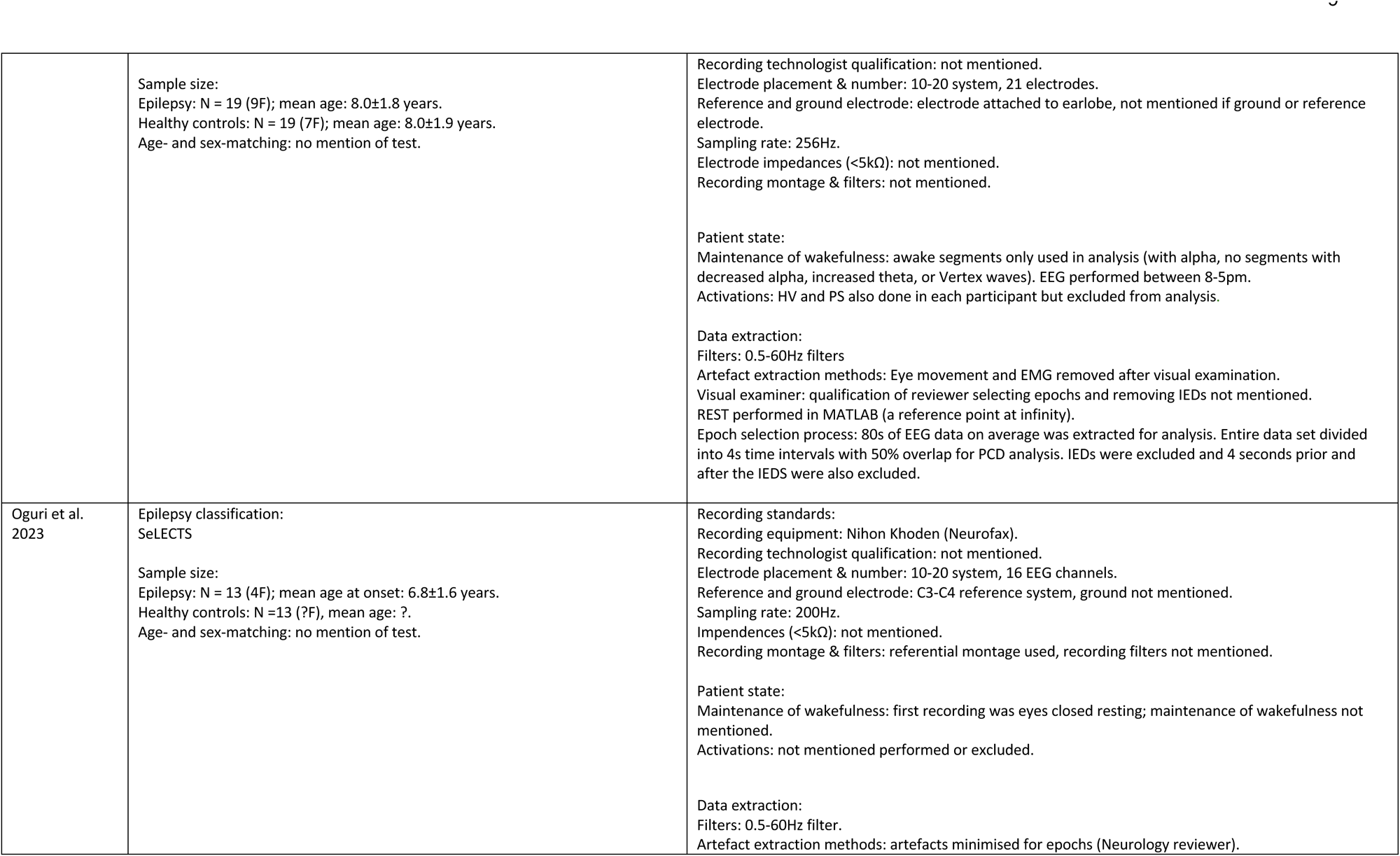
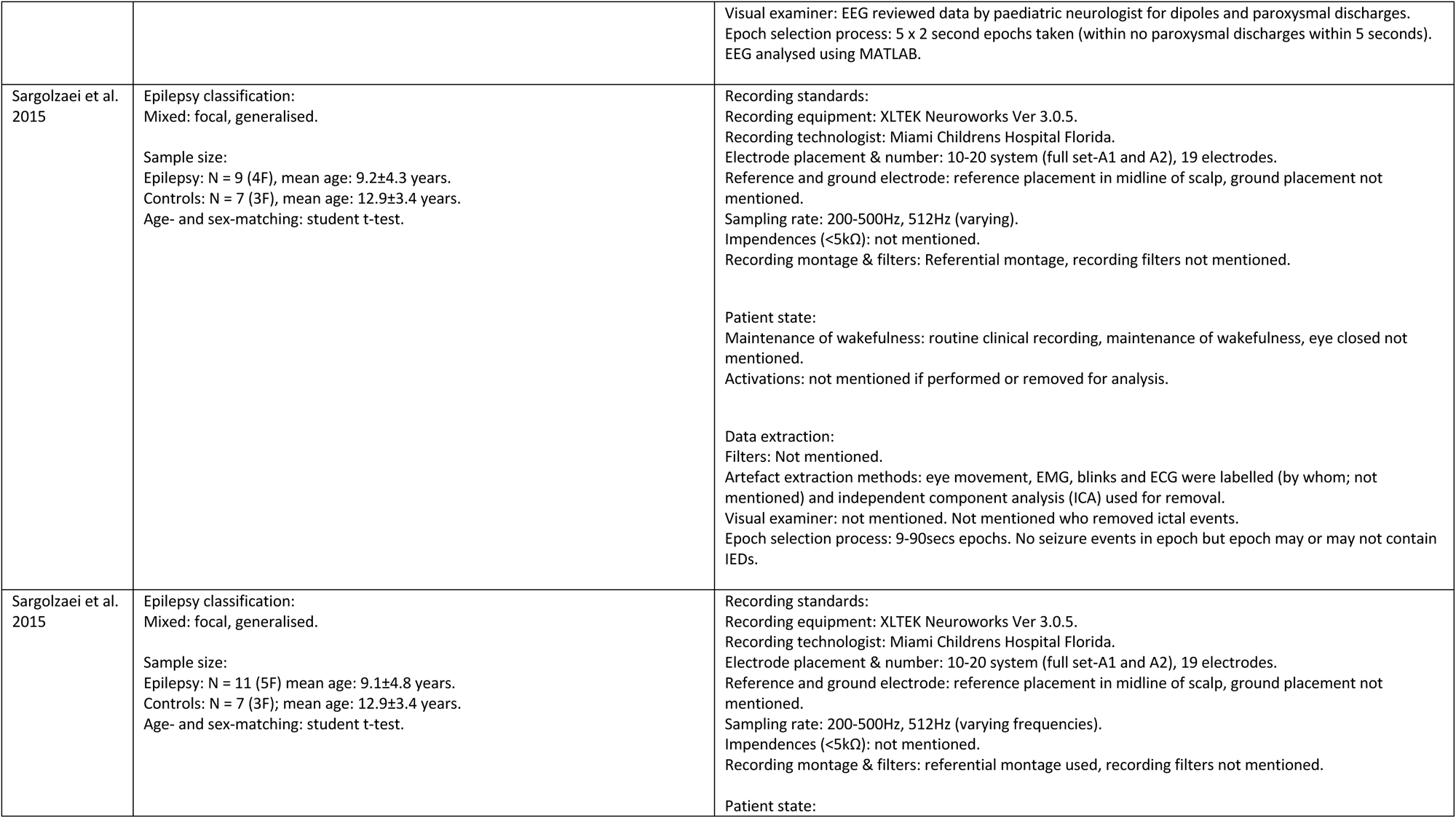
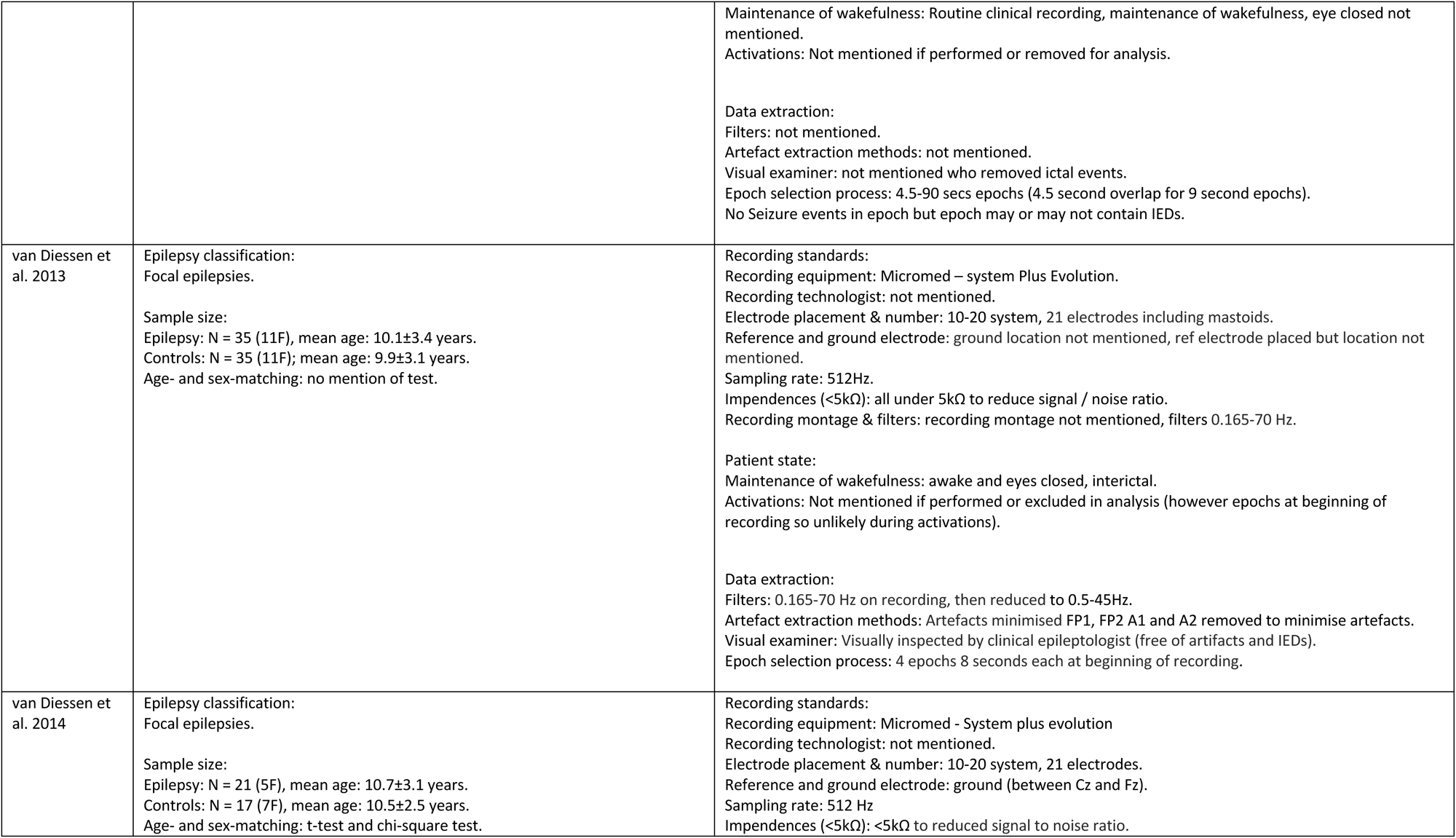
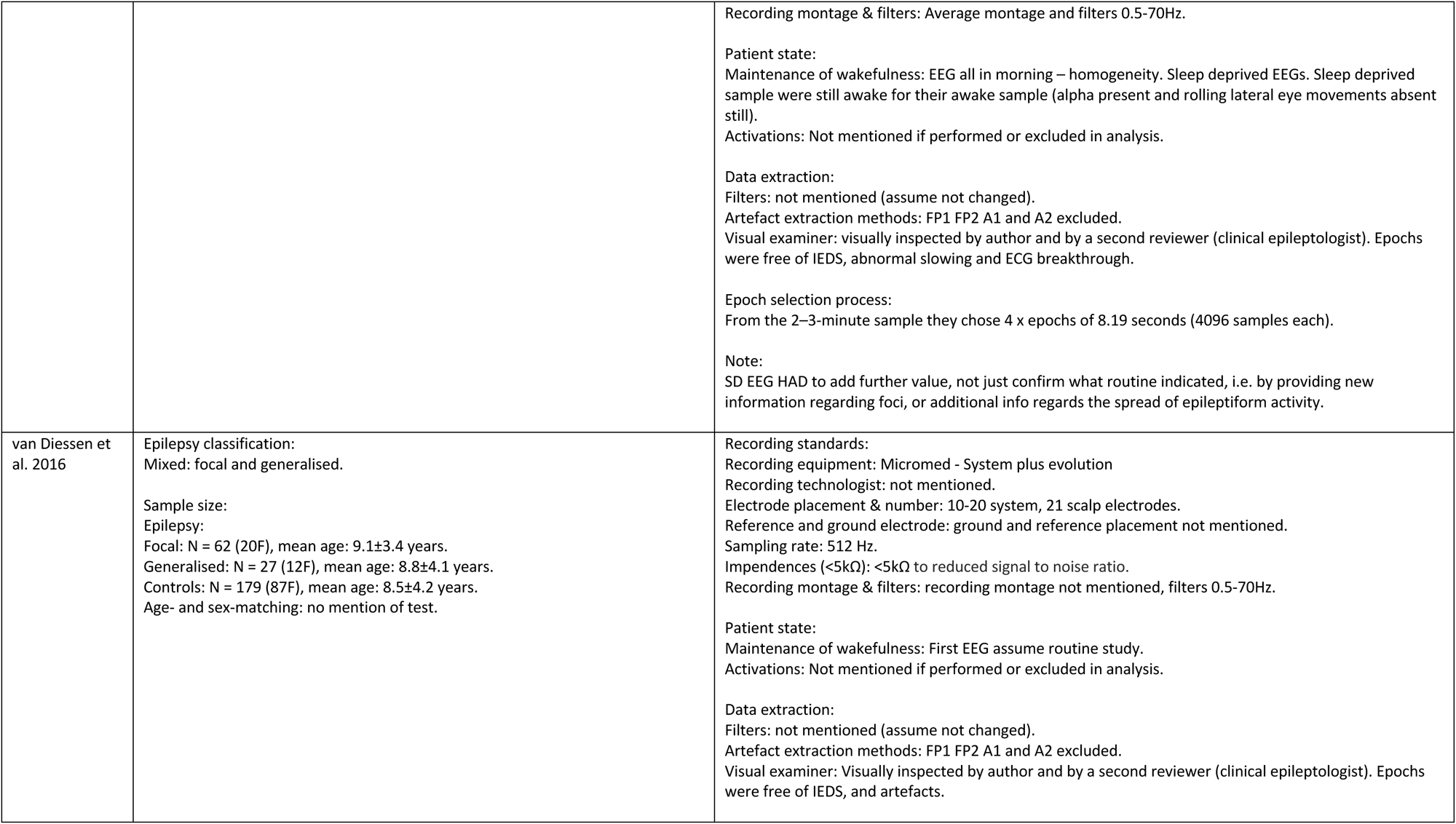
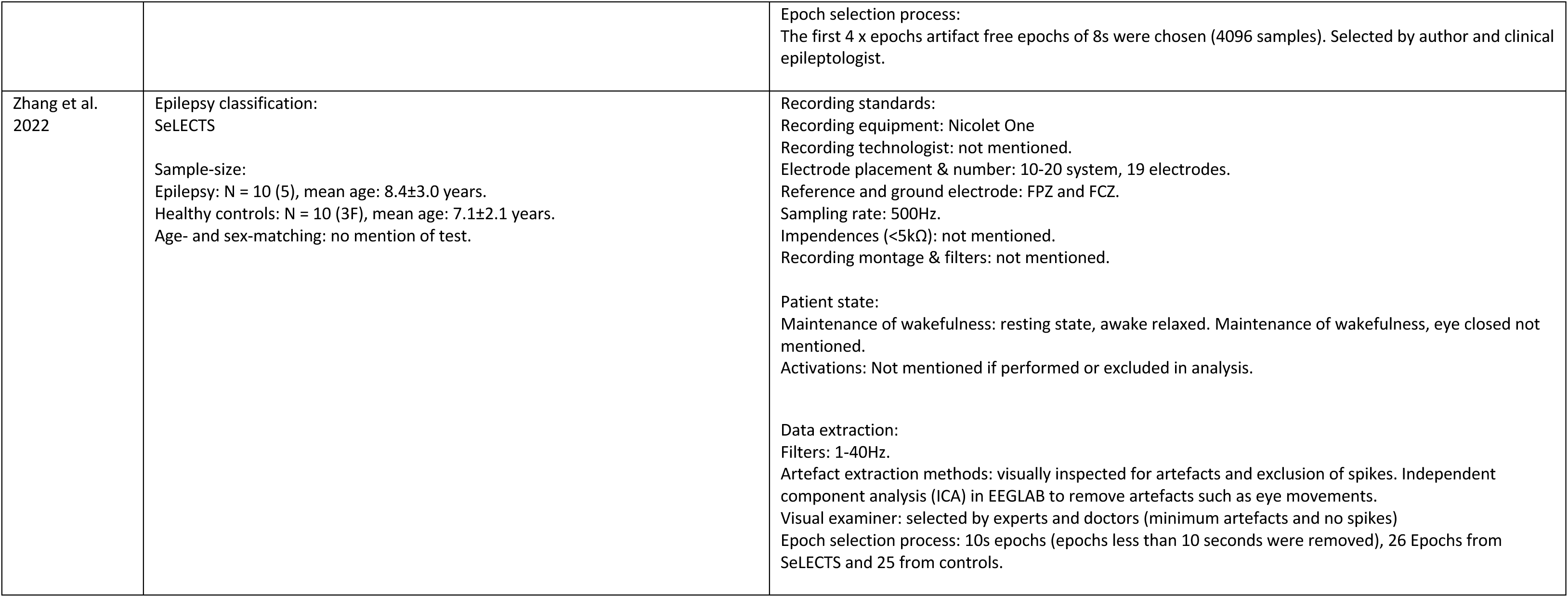
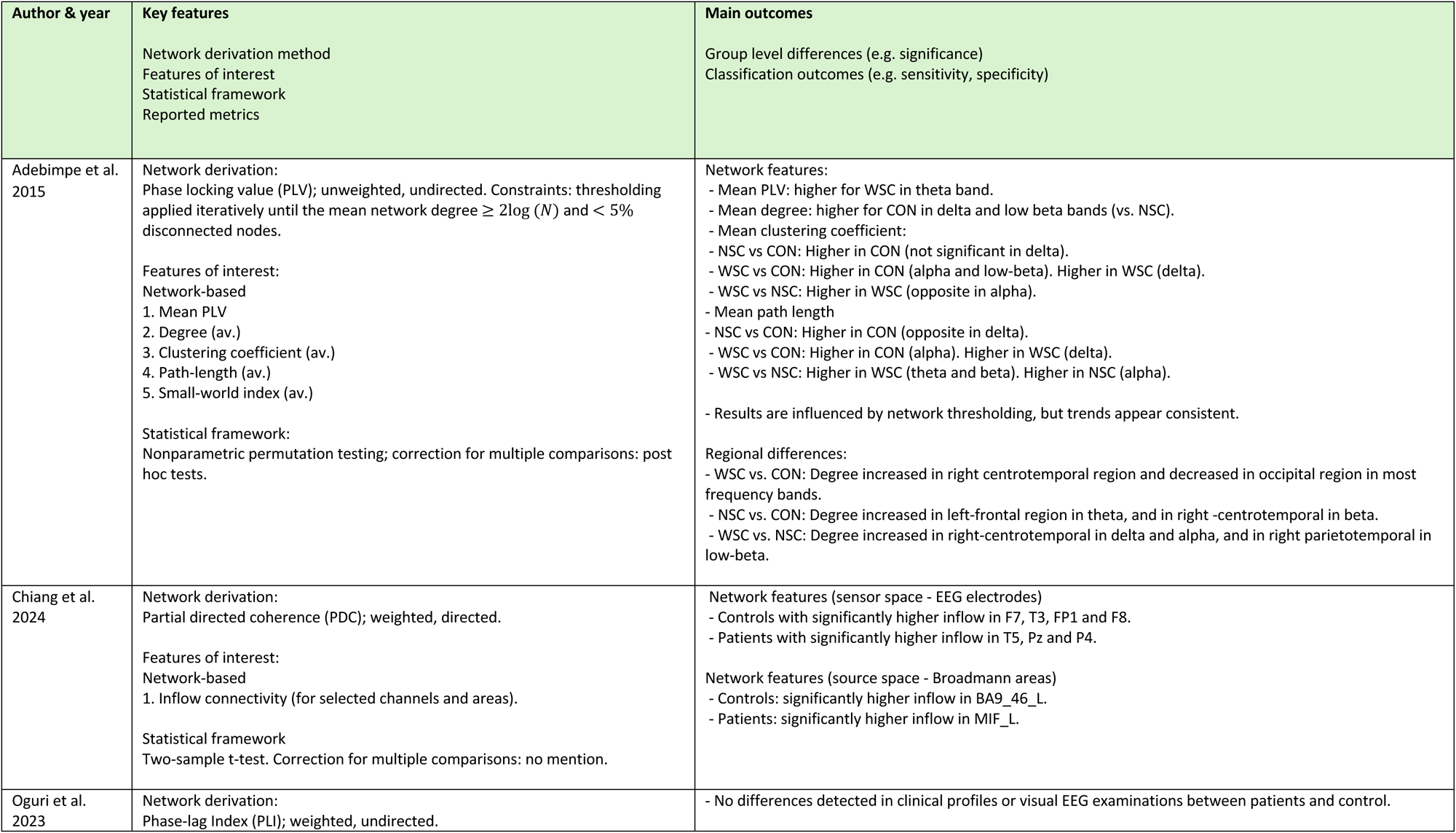
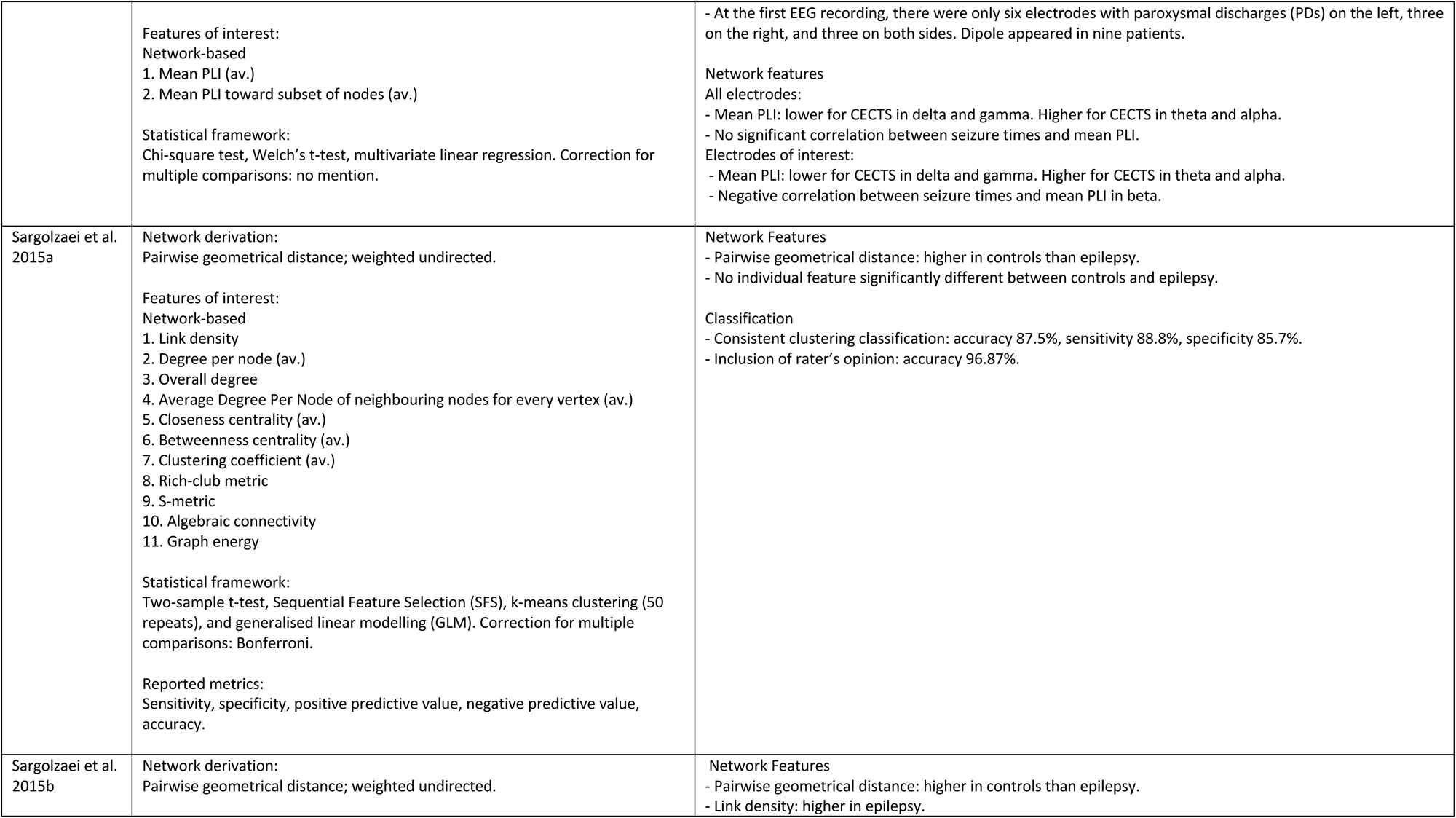
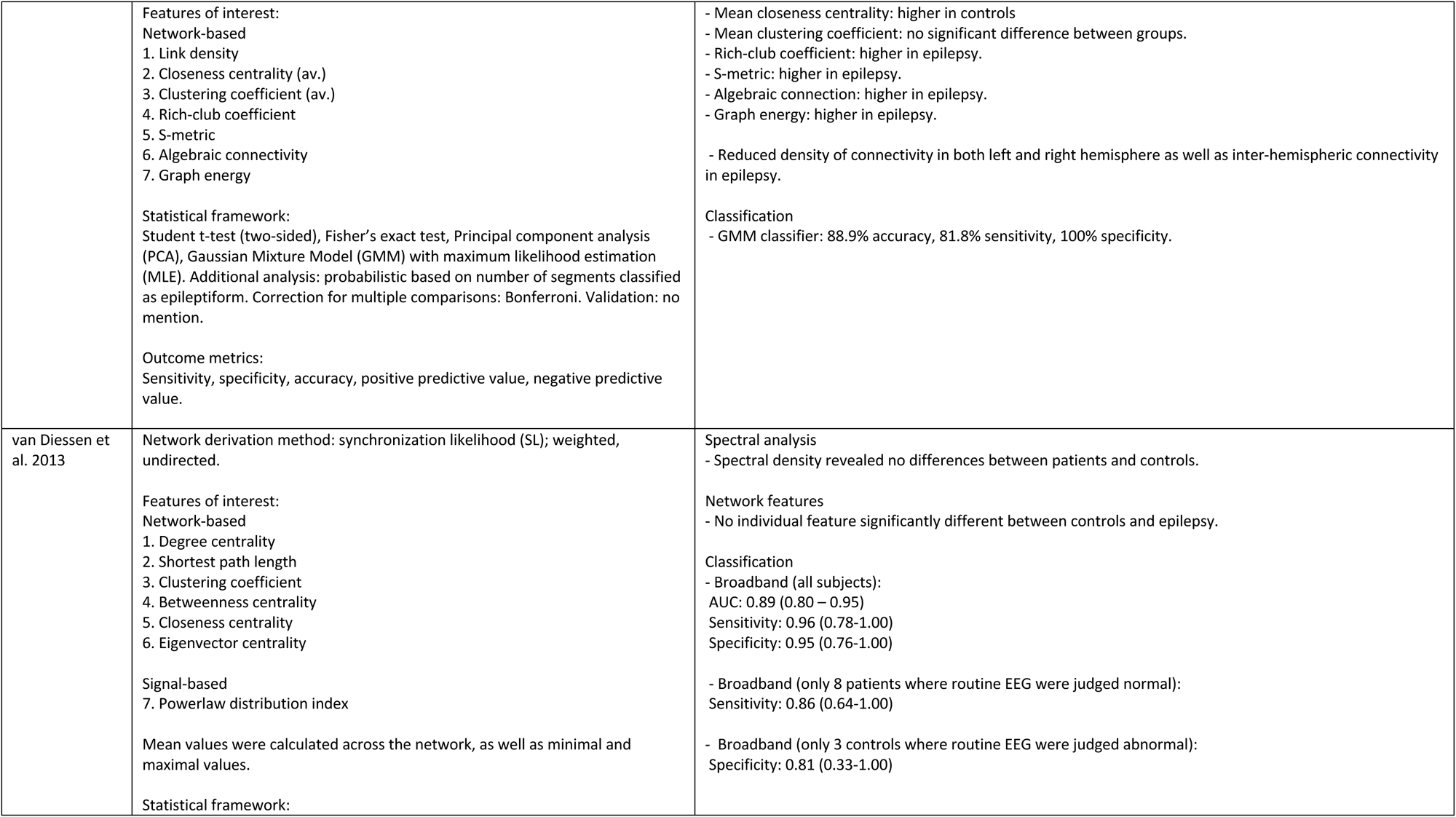
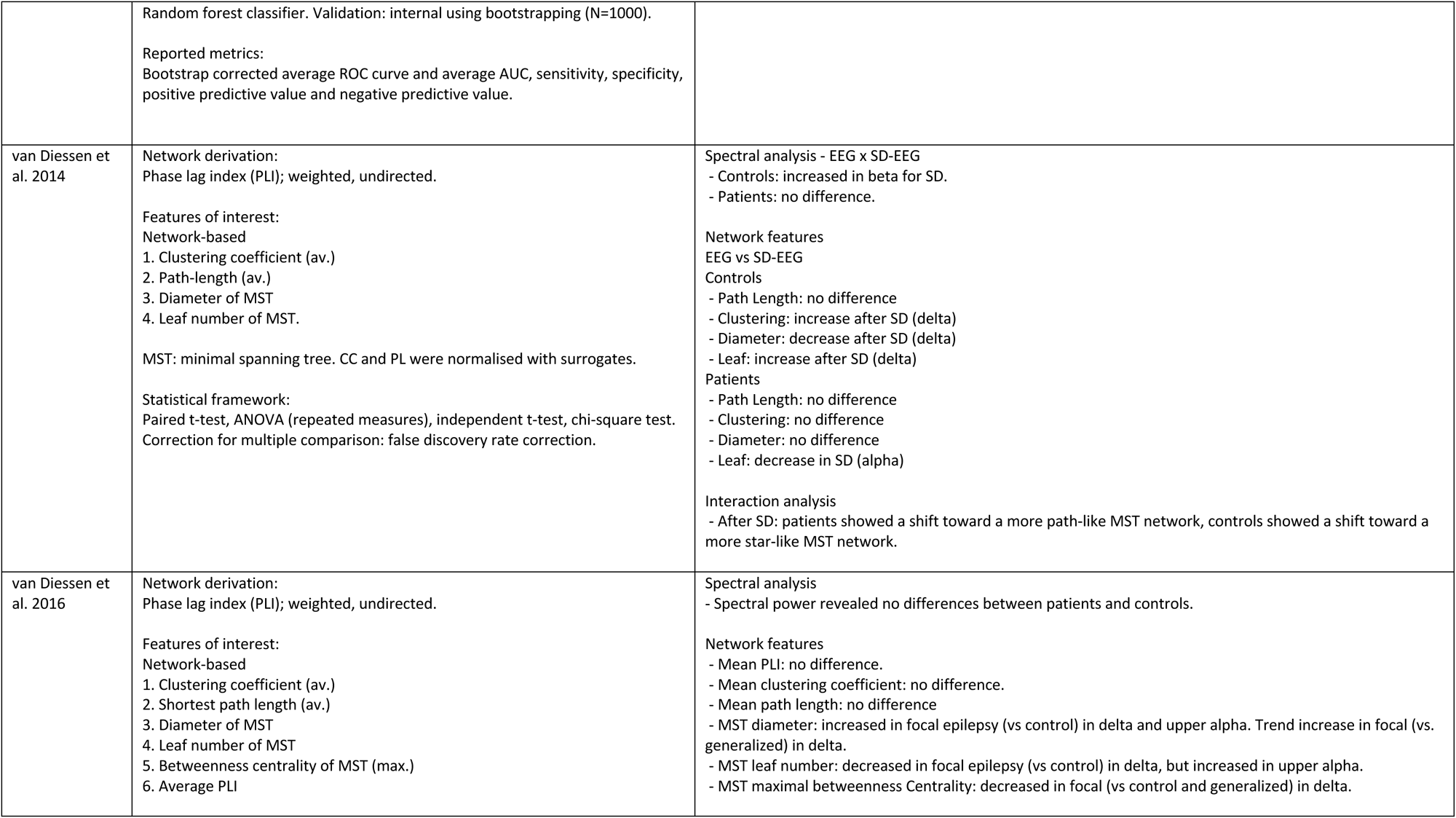
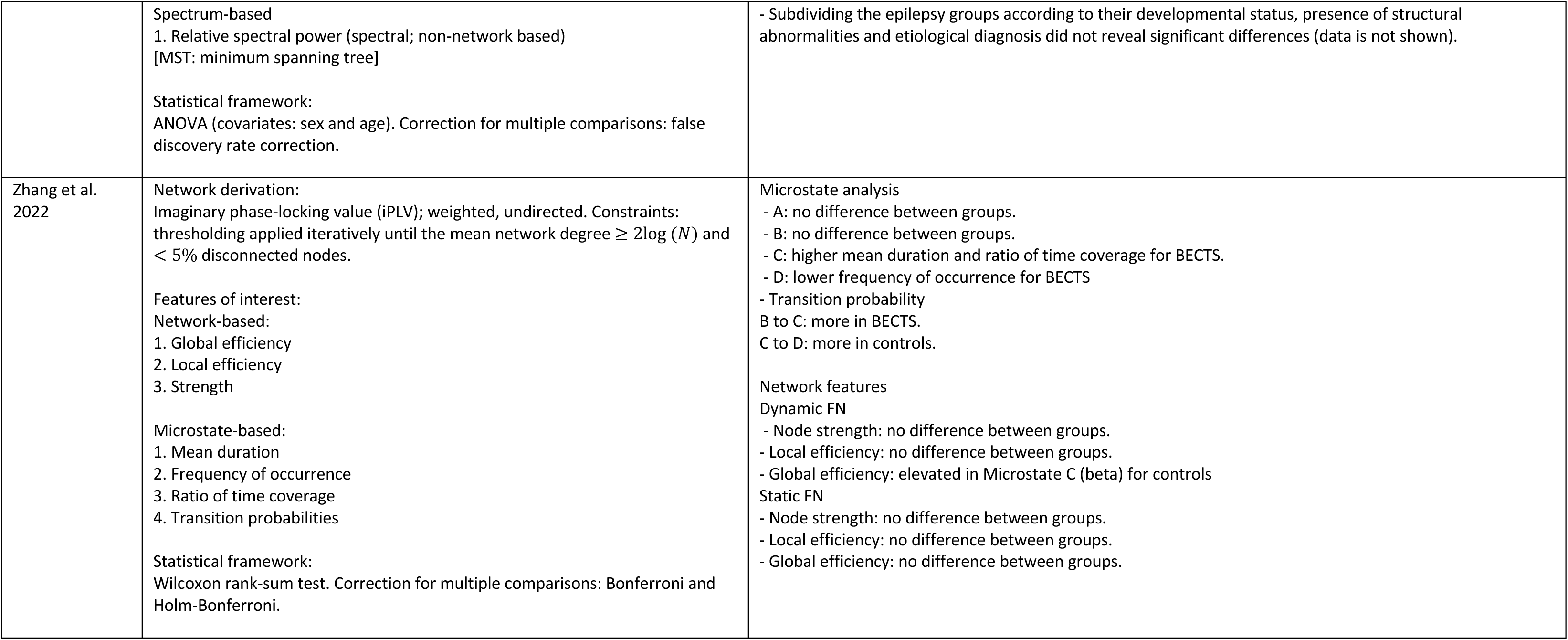
Details of the key PICO concepts extracted from the appraised nine papers. 3A: detailed description of the study population and EEG features; 3B: detailed descriptions of the key features and main outcomes. Note: the ILAE classification terminology for Self-limiting Epilepsy with Centro-Temporal Spikes (SeLECTS) has changed over the years. Previous terminology has included Benign Epilepsy with Centro-Temporal Spikes (BECTS) and Benign Rolandic Epilepsy (BRE). Depending on the year of publication the authors have used the current term reflecting this classification, however SeLECTS will be current term used throughout this document for ease of the reader.

### 3.2 Patient Characteristics

Four papers [16, 17, 18, 19] of the nine studies reviewed chose the SeLECTS epilepsy group to study. There were two studies that focused specifically on focal epilepsy [20, 21], with the remainder comprising of a mixed epilepsy group as their focus [22, 23, 24]. The majority of studies appraised were with relatively small cohort numbers (n<25 with matched control numbers), with the exception of two larger studies (n=89 in [24] & n=35 in [20]).

Healthy controls were equally matched against the study cohort in eight of the nine studies. The gender of the participants was matched with the exception of one study, which lacked information on age and sex [18]. Robust statistical analysis of age and sex matching was performed in one study [21]. Two other studies performed statistical analysis to demonstrate age and sex matching, and this showed that the difference between the age groups was not significant (p<0.05) [22, 23]. However, although the difference between the age groups was not significant (p<0.05), there was a strong trend (p< 0.086) which suggests it was not matched. This would be an important observation in certain childhood epilepsy cohorts with specific seizure types and narrow age of onset ranges.

Control participants were usually children referred to clinics having had childhood epilepsy excluded from their differential diagnosis following clinical review, EEG, and other adjunctive tests. Three studies describe the process of healthy control selection, detailing exclusion of epilepsy as stated above [20, 21, 24]. In all three studies from van Diessen they describe also a one year follow up to further confirm that the exclusion of epilepsy as a differential diagnosis had remained stable [20, 21, 24]. This is a favourable point to note in the control selection as epilepsy is often not diagnosed from one event and often has a watch and wait period [25]. This also emphasises the importance of a suitable period of review for inclusion or exclusion. Two studies detail that there was no family history of epilepsy, or comorbidities present in the control groups [17, 18]. This helps to limit the heterogeneous nature of the control group to reduce the confounders that could otherwise affect the results.

Associated comorbidities in the epilepsy group (both neurological and psychiatric) were considered in five of the nine studies [16, 17, 18, 20, 21] and these cases were excluded from the final epilepsy case cohort selection. Three of the nine studies did not mention if comorbidities were considered or excluded in the epilepsy study group [19, 22, 23], and one study specifically included neurological and psychiatric comorbidities but excluded epileptic encephalopathies [21]. Comorbidities are not uncommon in epilepsy but could reflect differing underlying mechanisms [26].

A history of febrile seizures was also considered an exclusion by one of the studies [20]. None of the other studies made any mention any febrile seizure exclusion. Febrile seizures typically occur in children aged six months to five years and occur with a fever greater than 38°C. Febrile seizures occur in 2-5% of children and although are considered benign, are associated with an increased risk of epilepsy [27]. Around 40% of medically refractory temporal lobe epilepsy and hippocampal sclerosis/ mesial temporal sclerosis on neuroimaging have a history of febrile seizures [28].

Post ictal effects and the exclusion of seizures in the days prior to collecting the EEG was considered in two papers [17, 20]. Post ictal states are defined as “transit abnormal neurological deficit or psychiatric symptoms following an epileptic seizure which is reflected on the EEG” [29]. This is an important variable to consider and exclude, due to the desynchronising nature of the EEG in the post ictal phase, which could potentially bias results.

Introduction of anti-seizure medications (ASM) and recurrent seizures would both introduce confounding effects on studies included for this review. It is therefore important to consider if the child with epilepsy was newly diagnosed and or taking ASMs at the time the EEG was taken. Three studies noted that the patient was newly diagnosed, this means the EEG would have been done early on in the diagnosis to limit the effect of recurrent seizures and the impact of ASMs [20, 21, 24]. Four studies did not mention the status of the patient and where in the diagnostic patient pathway the EEG was taken [17, 18, 22, 23]. One study noted the EEGs were specifically not from newly diagnosed patients [19]. The same study also detailed the epilepsy cohort were taking ASM treatment at the time of the EEG and were doing so for 32±24.6 months. Another study noted six of the 19 children of the epilepsy cohort were treated with ASMs [17]. The remaining studies removed the ASM cofounding factor from the epilepsy study group and all participants were medication naive.

The diagnostic criteria for the epilepsy diagnosis were only mentioned in three studies with two papers refer to the international league against epilepsy (ILAE) classifications for the diagnosis [17, 20, 24]. One study refers to “spikes” on the EEG as a diagnostic criterion [16], and one study mentioned centro-temporal spikes being activated in sleep as a diagnostic criterion for SeLECTS epilepsy cohort [18]. The remaining studies omit to refer to any clinical or electrographic diagnostic criteria proposed by ILAE that would form the basis of such diagnostic decisions.

### 3.3 EEG Recording characteristics

The minimal technical recording standards were in part mostly fulfilled across studies. Eight of the nine studies retrieved data from studies that used clinical grade EEG recording hardware and software that conformed to minimum recording standards set by professional bodies [30, 31]. It is unclear if the remaining study used recommended recording hardware due to them not stating the recording equipment used, but only that the Ant-Neuro software was used for dipole analysis [16].

Minimum recording standards set by ASET recommend the international 10-20 electrode placement is used with a minimum of 19 electrodes for full cortex coverage. Eight studies used to 10-20 system for recording the EEG. Seven of the eight studies conformed to the minimum electrode numbers except one study who stated 16 electrodes were used [18]. The 10-20 electrode placement system was used by all studies with exception of one study, who used the 10-10 system (and might potentially have used a cap) [16].

Minimum technical recording standards specify electrode impedances should be <5kΩ, and less than 2kΩ between electrodes [30, 31]. It is often difficult to achieve this with a cap particularly in the paediatric population as a high impedance will result in a high signal to noise ratio compromising the quality of the signal recorded. Three studies identify the importance of adhering to electrode impedances of <5kΩ [20, 21, 24].

Other recording parameters investigated by our review include the filters used and the recommended settings being between 0.5-70Hz thus allowing the optimal bandwidth of EEG frequencies to be recorded. Three studies mentioned these parameters however the other studies did not describe whether these parameters were adhered to [20, 21, 24]. Recorded montage and placement or reference and ground electrode are also noted only in part by half of the studies.

The patient state is an important factor when recording the EEG, particularly if a resting record is to be achieved, understanding how to achieve this is vital. It is typically challenging to collect artefact free or resting-state EEG from the paediatric population (particularly the younger age groups). Consequently, movement and eye movement artefact are frequently observed. Compliance increases however with age and maturity. Suitably qualified physiologists / technologists are expert in ensuring high quality recording standards are met, they are adept at artefact reduction (both biological and non-biological) and can identify ways to optimise signal quality of the EEG, especially during challenging recording sessions. However, qualifications of those performing the EEG were typically not reported. Three studies noted that for EEGs, subjects were awake with eyes are closed [20, 21]. One study further notes that wakefulness is monitored and ensures no drowsiness features occurred [17]. The remaining five did not document the potentially changing state of wakefulness of subjects during the EEG.

During routine EEG, it is standard to perform activations such as hyperventilation and photic stimulation. One study identified that this was a variable that would affect the background state of the EEG and removed that particular segment from analysis [17]. One study mentioned that only the beginning of the test was selected for analysis, thus suggesting that activations were not performed in this segment as it is usual for the recording physiologist / technologist to initially obtain a segment of resting background to consider the risk in performing the aforementioned activations [20]. The remaining seven studies did not provide details on whether activations were included or excluded from the epoch analysis. Activations may change the background EEG signals significantly. It is not uncommon for the EEG to increase in amplitude up to four-fold during hyperventilation, and background frequencies may show considerable slowing.

Upon obtaining data the pre-processing requires the adaptation of filters. The changes in filter settings were mentioned in three studies [16, 19, 20]. They outlined reducing high frequency filters from the recorded 70Hz to 30Hz, 45Hz, and 40 Hz respectively. Altering filters may change the signal considerably, the morphology of the waveforms will change as IEDs become rounded as the high frequency filter is reduce. Phase shifting some of the signals may also occur during adaptation of filters [32; p. 132].

Five studies provided brief descriptions on who undertook the identification of epileptiform abnormalities [16, 19, 20, 21, 24]. Independent component analysis (ICA) was used by two studies to remove artefacts [22, 23]. Ictal events were noted to be removed by all studies and IED removal was consistent in all epochs chosen in all studies with the exception of two who argued that cortical spikes could also be identified in the control cohort without them being considered epilepsy segments [22, 23]. This is a potential confounder as although IED are seen in a small percentage 0.3-0.5% of normal adult subjects the presence of IEDs is more likely in epilepsy group than the control group [33, 34].

Epoch lengths and quantity are shown in Table 4:

**Table 4.**

| Study | Epoch length (seconds) | Number of epochs per study group |
| --- | --- | --- |
| Adebimpe 2015 | 2 | 5 |
| Chiang 2024 | 80 | 4 |
| Oguri 2023 | 2 | 5 |
| Sargolzaei 2015 a | 9-90 | 3-77 depending on subject |
| Sargolzaei 2015 b | 4.5-90 | Not mentioned |
| van Diessen 2013 | 8 | 4 |
| van Diessen 2014 | 8.19 | 4 |
| van Diessen 2016 | 8 | 4 |
| Zhang 2022 | 10 | 26 |

Two studies considered removing 4 and 5 seconds respectively from pre and post IEDs, which was insightful to remove any potential network changes that have occurred pre and post any paroxysmal activity [17, 18]. Frequency band definition was commonly reported, however, overlapping frequencies bands was observed in several studies.

### 3.4 Network-based analysis

Functional networks were derived from EEG segments with different techniques by the reviewed studies. Five of the nine studies were phase-based (e.g. Phase Locking Value, (imaginary) Phase Lag Index; [16, 18, 19, 21, 24]. The other four studies used amplitude-based methods [17, 20, 22, 23]. Certain network derivation techniques such as the synchronization likelihood were developed to identify linear and nonlinear correlation or interactions [20].

The majority of the studies (7/9) derived features of interest from weighted, undirected functional networks [18, 19, 20, 21, 22, 23, 24]; one study focused on unweighted, undirected networks [16], and one study on weighted, directed networks [17]. Two studies described a dynamic thresholding procedure during network construction [16, 19].

A primary focus of the review was the evidence reported by the studies about the potential of network-based features to differentiate between a cohort of participants with childhood epilepsies and a cohort of controls. There were several significant differences amongst the studies, a wide range of candidate features (see Table 5) and outcome measures were used. A comprehensive summary of observed group-level differences for different graph measures across several frequency-bands is provided in Table 5.

**Table 5.**

| Network measure | Delta | Theta | Alpha | Beta | Gamma | Broad band |
| --- | --- | --- | --- | --- | --- | --- |
| Mean node strength | 3, 1, 8 | 1*, 3, 8 | 3, 1, 8 | 3*, 1, 8 | 3 | 5 |
| Mean degree | 1** | 1 | 1 | 1** |  | 4 |
| Mean clustering coefficient | 1*, 8 | 1**, 8 | 1, 8 | 1***, 8 |  | 6, 4, 5 |
| Mean path length | 1, 8 | 1**, 8 | 1, 8 | 1**, 8 |  |  |
| FCN pairwise distance |  |  |  |  |  | 4, 5 |
| Link density |  |  |  |  |  | 4, 5 |
| Average degree per node of neighbouring nodes for every vertex |  |  |  |  |  | 4 |
| Rich club metric |  |  |  |  |  | 4, 5 |
| S-metric |  |  |  |  |  | 4, 5 |
| Algebraic connectivity |  |  |  |  |  | 4, 5 |
| Graph energy |  |  |  |  |  | 4, 5 |
| Betweenness centrality |  |  |  |  |  | 6, 4 |
| Closeness centrality |  |  |  |  |  | 6, 4, 5 |
| Eigenvector centrality |  |  |  |  |  | 6 |
| Degree centrality |  |  |  |  |  | 6 |
| Shortest path length |  |  |  |  |  | 6 |
| MST diameter | 8 | 8 | 8 | 8 |  |  |
| MST leaf number | 8 | 8 | 8 | 8 |  |  |
| MST BC (max.) | 8* | 8 | 8 | 8 |  |  |

Blue: greater in controls, Red: greater in epilepsy, Black: not significant. 1\*: only controls vs. epilepsy epochs with spikes; 1\*\*: only controls vs. epilepsy epochs without spikes; 1\*\*\*: only low-beta (13.5Hz-20Hz); 3\*: trend, but not significant.; 8\*: only higher in controls compared to focal (not generalized).

Outcome measures for the classification-based studies included sensitivity, specificity, and accuracy [20, 22, 23]. One study also reported the area under the ROC-curve (AUROC) as well as confidence intervals for the reported outcome measures [20], whereas two studies reported positive and negative predictive values [22, 23]. One study combined an expert rater with the classification model [22], which resulted in increased overall performance in terms of accuracy. One study provided a probabilistic score as well as binary predictions [23].

The majority of the studies applied types of correction for multiple comparisons (e.g. Bonferroni or false-discovery rate). However, none of the studies included an explicit sample-size calculation or calculations on the appropriate number of potential features given the sample-size.

Given the heterogeneity and diversity between the studies at different stages of the analysis pipeline – differences in epochs, pre-processing steps, network derivation method, features of interest, statistical frameworks, and outcome metrics – the combined or compounded heterogeneity is significant.

## 4. Discussion

### 4.1 Summary of findings

The review identified a total of nine studies that explored network-based abnormalities in childhood epilepsies from EEG. The vast majority of these studies used routine scalp EEG, with some using high-density caps. Most studies included a control cohort consisting of age- and gender-matched subjects that were otherwise healthy. Some studies included one specific type or syndrome of epilepsy within the epilepsy cohort (e.g. SeLECTS), however most studies explored network abnormalities within a broader cohort (e.g. focal epilepsies or a mix of focal and generalised epilepsies). Age varied across studies, with different mean ages (primarily < 14 years) and standard deviation typically ranging from 2-4 years (with the exception of one study: 0.2 years). The studies examined a variety of different candidate features and outcome measures (e.g. statistical framework). For example, there was a high degree of variability in the type and duration of epochs used for network-derivation, the type of network-derivation techniques (e.g. linear, non-linear, amplitude- and phase-based), and the candidate graph measures. In terms of outcome measures, the studies can be divided in group-level tests (e.g. by using ANOVA) and classifier-based models (e.g. by using a decision tree). Group-level tests were usually assessed by significance, but not always corrected for multiple comparisons or presented with a confidence interval. Classifier-based approaches assessed the performance of the final model with standard metrics such as the sensitivity, specificity, and accuracy, but at times without specific details on appropriate validation (e.g. hold-out or cross-validation) or avoidance of overfitting (e.g. sample-size calculations).

A common limitation is the lack of standardised reporting, e.g. covering the critical steps included in the relevant guidelines from the EQUATOR network. However, despite the significant differences in methodology and classification frameworks, the studies present result greater than chance across a subset of features, which suggests that network-based analysis of EEG in childhood epilepsy could potentially lead to novel biomarkers for epilepsy and seizures (i.e. towards Phase II or Phase III evidence).

### 4.2 Strengths and weaknesses of the studies

The majority of the studies did not include specific discussion of the strengths of their study. Several study design components were identified by the reviewers as strengths, including assessing the impact of comorbidities [16, 17, 18, 20, 21], monitoring of wakefulness [17], removal of activation [17], and reported the use of conservative statistical methods such as correcting for multiple comparisons [16, 19, 21, 22, 23, 24].

The most prevalent and significant methodological weakness identified in the studies was the small sample size. Only two studies had sample size greater then 25 in each subpopulation (control and epilepsy), and the majority included cohorts with fewer than 15. This significantly impacted the potential to carry out any subsequently subgroup analysis (e.g. impact of age, or specific syndromes). Several studies were also impacted by a lack of clarity in epoch and feature selection, statistical framework (e.g. age- and gender-matching), and validation of classification paradigm. Taken together, this significantly impacts how well the results can be interpreted and compared, as it limits the generalizability and potential robustness of the results. Examples of reported limitations include: inability to carry out further subgroup analysis due to small sample-size, such as comparing the measures syndromes [17, 20, 24]; studying the relationship between network connectivity and cognitive function [17], assessing the impact of seizure-frequency [18]; inaccuracy of networks inferred from scalp EEG as an accurate mechanistic representation of the underlying cortical (i.e. anatomical) network [16]; inability to explore difficulty in collecting artefact-free, resting-state EEG in very young children [20]; potential lack of specificity or generalizability related to the control-group [21], potential impact of confounding variables, such as ASM [21] or cognitive state [19].

### 4.3 Strengths and weaknesses of the review

This review is to our knowledge the first to provide a comprehensive overview of network-based abnormalities from EEG in childhood epilepsies. A strength is that the review was conducted by multiple authors independently of each other whose expertise spanned epilepsy (including EEG data collection and analysis), network analysis (in both adult and paediatric cohorts) and digital health. A weakness of the review is that the limited number of studies combined with significant heterogeneity in study design (e.g. epochs, network derivation) and population (e.g. age groups, epilepsy syndrome) hampers a rigorous comparison between the studies and their approaches, and consequently their overall benefits and potential limitations. Another weakness is that this review did not identify two studies that met our inclusion criteria that were appraised in an earlier systematic review that focussed on idiopathic generalized epilepsies [12, 35, 36]. These studies used correlation-based analysis between the time-series of different channels (i.e. equivalent to node strength). This is not a network analysis in the traditional sense, which might explain why the current search strategy failed to identify them. However, the overall findings of these studies are consistent with the presented results in terms of main outcomes and subsequent meta-analysis.

The search was designed to identify candidate network-based biomarkers in EEG; however, several studies reviewed at the full-text screening state examined the EEG using neural networks, which is a branch of machine learning where a neural net learns to handle specifically set tasks (e.g. classifying between two distinct groups). After discussion, the authors concluded to exclude these studies if they did not include an explicit derivation of (functional) network structures in constructing candidate features.

### 4.4 Conclusion and future research

This review provides an overview of different types of studies attempting to identify abnormalities in childhood epilepsies using network-analysis of EEG. These studies can roughly be divided in those that assess group-level differences (e.g. by means of a particular statistical test) or those that utilise a classifier-based approach (e.g. by means of a particular statistical or machine-learning model). Whereas several studies examined network-differences for a specific type of childhood epilepsy (e.g. SeLECTS), other studies used focal epilepsies as a group or a mix of focal and generalised epilepsies.

A key focus for future research should be to increase overall sample-sizes. Future work should adhere to the relevant guidelines developed by the Equator Network, such as the STARD or STROBE guidelines, which would enable more rigorous comparisons between studies, and consequently a better foundation for assessing the potential of network-based approaches in developing novel biomarkers for childhood epilepsies. This would potentially also stimulate future research that would explore how these network-based features vary over time or specific ages, and whether specific statistical models need to be developed that would facilitate this.

## Author contributions

Design of the review: all authors. AW conducted the database searches. KM conducted the searches and initial screening. KM, WW and LJ conducted the title, abstract and full text screening and data extraction. KM, WW and LJ drafted the first version of the review and all authors contributed revisions.

## Disclaimer

This work was commissioned and funded by SBRI Healthcare. SBRI Healthcare is an Accelerated Access Collaborative (AAC) initiative, in partnership with the Health Innovation Network. The views expressed in the publication are those of the author(s) and not necessarily those of SBRI Healthcare or its stakeholders.

## Data Availability

All data produced in the present work are contained in the manuscript

