## Supplementary Materials for "Network-based biomarkers in background electroencephalography in childhood epilepsies – A scoping review and narrative synthesis"

a) University Hospital Southampton NHS Foundation Trust, Southampton, United Kingdom
b) Neuronostics, Bristol, United Kingdom
c) Centre for Systems Modelling and Quantitative Biomedicine, University of Birmingham, Birmingham B15 2TT, United Kingdom
d) Institute of Metabolism and Systems Research, University of Birmingham, Birmingham B15 2TT, United Kingdom
e) Cornwall Health Library, Truro, United Kingdom
f) Cornwall Partnership NHS Foundation Trust, Bodmin, United Kingdom
g) University of Plymouth, Plymouth, United Kingdom

*: joint first authors
†: joint senior authors

**A: Table SM-1:** PRISMA-ScR checklist adapted from PRISMA Extension for Scoping Reviews (PRISMA-ScR): Checklist and Explanation | Annals of Internal Medicine (acpjournals.org)

*Table.* PRISMA-ScR Checklist

| Section | Item | PRISMA-ScR Checklist Item |
|---|---|---|
| **Title** | 1 | Identify the report as a scoping review. |
| **Abstract** | | |
| Structured summary | 2 | Provide a structured summary that includes (as applicable) background, objectives, eligibility criteria, sources of evidence, charting methods, results, and conclusions that relate to the review questions and objectives. |
| **Introduction** | | |
| Rationale | 3 | Describe the rationale for the review in the context of what is already known. Explain why the review questions/objectives lend themselves to a scoping review approach. |
| Objectives | 4 | Provide an explicit statement of the questions and objectives being addressed with reference to their key elements (e.g., population or participants, concepts, and context) or other relevant key elements used to conceptualize the review questions and/or objectives. |
| **Methods** | | |
| Protocol and registration | 5 | Indicate whether a review protocol exists; state if and where it can be accessed (e.g., a Web address); and if available, provide registration information, including the registration number. |
| Eligibility criteria | 6 | Specify characteristics of the sources of evidence used as eligibility criteria (e.g., years considered, language, and publication status), and provide a rationale. |
| Information sources* | 7 | Describe all information sources in the search (e.g., databases with dates of coverage and contact with authors to identify additional sources), as well as the date the most recent search was executed. |
| Search | 8 | Present the full electronic search strategy for at least 1 database, including any limits used, such that it could be repeated. |
| Selection of sources of evidence† | 9 | State the process for selecting sources of evidence (i.e., screening and eligibility) included in the scoping review. |
| Data charting process‡ | 10 | Describe the methods of charting data from the included sources of evidence (e.g., calibrated forms or forms that have been tested by the team before their use, and whether data charting was done independently or in duplicate) and any processes for obtaining and confirming data from investigators. |
| Data items | 11 | List and define all variables for which data were sought and any assumptions and simplifications made. |
| Critical appraisal of individual sources of evidence§ | 12 | If done, provide a rationale for conducting a critical appraisal of included sources of evidence; describe the methods used and how this information was used in any data synthesis (if appropriate). |
| Summary measures | 13 | Not applicable for scoping reviews. |
| Synthesis of results | 14 | Describe the methods of handling and summarizing the data that were charted. |
| Risk of bias across studies | 15 | Not applicable for scoping reviews. |
| Additional analyses | 16 | Not applicable for scoping reviews. |
| **Results** | | |
| Selection of sources of evidence | 17 | Give numbers of sources of evidence screened, assessed for eligibility, and included in the review, with reasons for exclusions at each stage, ideally using a flow diagram. |
| Characteristics of sources of evidence | 18 | For each source of evidence, present characteristics for which data were charted and provide the citations. |
| Critical appraisal within sources of evidence | 19 | If done, present data on critical appraisal of included sources of evidence (see item 12). |
| Results of individual sources of evidence | 20 | For each included source of evidence, present the relevant data that were charted that relate to the review questions and objectives. |
| Synthesis of results | 21 | Summarize and/or present the charting results as they relate to the review questions and objectives. |
| Risk of bias across studies | 22 | Not applicable for scoping reviews. |
| Additional analyses | 23 | Not applicable for scoping reviews. |
| **Discussion** | | |
| Summary of evidence | 24 | Summarize the main results (including an overview of concepts, themes, and types of evidence available), link to the review questions and objectives, and consider the relevance to key groups. |
| Limitations | 25 | Discuss the limitations of the scoping review process. |
| Conclusions | 26 | Provide a general interpretation of the results with respect to the review questions and objectives, as well as potential implications and/or next steps. |
| **Funding** | 27 | Describe sources of funding for the included sources of evidence, as well as sources of funding for the scoping review. Describe the role of the funders of the scoping review. |

JBI = Joanna Briggs Institute; PRISMA-ScR = Preferred Reporting Items for Systematic reviews and Meta-Analyses extension for Scoping Reviews.
* Where *sources of evidence* (see second footnote) are compiled from, such as bibliographic databases, social media platforms, and Web sites.
† A more inclusive/heterogeneous term used to account for the different types of evidence or data sources (e.g., quantitative and/or qualitative research, expert opinion, and policy documents) that may be eligible in a scoping review as opposed to only studies. This is not to be confused with *information sources* (see first footnote).
‡ The frameworks by Arksey and O'Malley (6) and Levac and colleagues (7) and the JBI guidance (4, 5) refer to the process of data extraction in a scoping review as data charting.
§ The process of systematically examining research evidence to assess its validity, results, and relevance before using it to inform a decision. This term is used for items 12 and 19 instead of "risk of bias" (which is more applicable to systematic reviews of interventions) to include and acknowledge the various sources of evidence that may be used in a scoping review (e.g., quantitative and/or qualitative research, expert opinion, and policy documents).

### B: Example Search String

1 (epilep* or seizure* or convuls*).ab,kf,ti. 358792
2 exp epilepsy/ 278254
3 exp seizure/ 223943
4 1 or 2 or 3 467329
5 "toddler*".ab,kf,ti. 19733
6 toddler/ 6921
7 "child*".ab,kf,ti. 2167956
8 exp childhood/ 110322
9 "p?ediatric*".ab,kf,ti. 770243
10 pediatrics/ 94536
11 juvenile*.ab,kf,ti. 119664
12 juvenile/ 56579
13 "youth*".ab,kf,ti. 129304
14 "adolescen*".ab,kf,ti. 490371
15 exp adolescence/ 95878
16 "teen*".ab,kf,ti. 51189
17 (young people or young person*).ab,kf,ti. 57381
18 5 or 6 or 7 or 8 or 9 or 10 or 11 or 12 or 13 or 14 or 15 or 16 or 17 2942184
19 4 and 18 104454
20 Panayiotopoulos syndrome.ab,kf,ti. 269
21 exp childhood epilepsy/ 7408 (this includes SH terms for the specific types of childhood epilepsy)
22 20 or 21 7462
23 19 or 22 105797
24 EEG.ab,kf,ti. 146431
25 electroencephalogram.ab,kf,ti. 31327
26 electro-encephalogram.ab,kf,ti. 296
27 exp electroencephalogram/ 166498
28 MEG.ab,kf,ti. 13884
29 "Magnetoencephalogra*".ab,kf,ti. 11794
30 "Magneto-encephalogra*".ab,kf,ti. 320
31 magnetoencephalography/ 14901
32 24 or 25 or 26 or 27 or 28 or 29 or 30 or 31 243204
33 network*.ab,kf,ti. 907398
34 exp nerve cell network/ 85735 (this includes the SH term ‘resting state network’)
35 33 or 34 931871
36 23 and 32 and 35 929 (with network free text term and nerve cell network SH term)
37 23 and 32 20718 (without any network terms)
